# Systematic Review Search Strategy Development Tools: A Practical Guide for Expert Searchers

**DOI:** 10.1101/2025.07.10.25331296

**Authors:** Robin A. Paynter, Gaelen P. Adam, Allison Hedden-Gross, Claire Twose, Christiane Voisin

## Abstract

**Objectives:** To provide the expert searcher community with reliable reviews on systematic review search strategy development tools (SSDT). We conducted a descriptive comparative analysis of the free SSDT listed on the <u>Systematic Review Toolbox</u>^1–2^ (SR Toolbox) website.

**Study Design and Setting:** Our search expert panel is composed of five biomedical librarians currently active in the <u>US Agency for Healthcare Research and Quality (AHRQ) Effective</u> <u>Health Care Program Evidence-based Practice Centers (EPC)</u>. Two panel members reviewed each included SSDT that met inclusion criteria with any assessment disagreements resolved via consensus. We created a table of included SSDT, categorized them by search development step, rated their performance across our key domains and added narrative reviews outlining how to use them.

**Results:** Of the 102 total search tools listed on the SR Toolbox search software webpage (initial search September 2021 N=78; update search February 2023 N=24 new tools), we determined 21 were *search strategy development tools* and eligible for review. We found the preponderance of the 21 included SSDT are designed for the initial strategy development steps (i.e., scoping, keyword/phrase and subject term discovery), though there are tools in most steps meeting minimum performance criteria, with the notable exception of strategy logic evaluation. Our reviews include a graphical table covering key performance summary ratings and a narrative section providing more detailed information.

**Conclusion:** We created a quick and intuitive SSDT guide for expert searchers that is organized by search strategy development phase, evaluates key features, and provides advice on how to use the tool effectively.

## 1. INTRODUCTION

The continuing rapid growth in the international medical research literature has increased both the complexity and time demands for librarians/information specialists attempting to identify studies for a systematic review (SR). At the same time, technological innovation has led to the development of machine-learning, text-mining, artificial intelligence, and other software tools applicable to several processes involved in SRs, with many intended to help with search processes.

Tools developed to assist in SR processes are catalogued in the <u>Systematic Review Toolbox</u>^1–2^, hereafter “SR Toolbox.” The SR Toolbox is an online catalogue of SR methods guidance sites as well as software tools supporting SR processes. Since 2014, the SR Toolbox has been edited and maintained by a team from the United Kingdom. This team includes researchers from the York Health Economics Consortium (University of York), the Evidence Synthesis Group at Newcastle University/NIHR Innovation Observatory, and the School of Health and Related Research at the University of Sheffield. The recent growth in search software tools, however, has not been accompanied by any resource providing expert searchers reviews of the tools themselves.

The objective of this project therefore was to create a review resource for expert searchers to quickly and easily identify useful tools for search strategy development (SSDT). We conducted a descriptive comparative analysis of the free SSDT listed on the SR Toolbox website.

## 2. METHODS

Our team of investigators was composed of five search expert biomedical librarians with 60+ years of combined SR searching experience for the <u>AHRQ Effective Healthcare Program</u> <u>Evidence-based Practice Centers</u> (EPC). Beginning in September 2021, we identified and reviewed 21 SSDT available and free on SR Toolbox. We chose the SR Toolbox because it is the most comprehensive and actively updated list of software dedicated to the conduct of SRs. We focused on reviewing SSDT for a search expert audience who develop, perform, document, and deliver literature searches for biomedical and health-related evidence synthesis products, e.g., SRs, scoping reviews, living reviews, etc.

We categorized the 21 SSDT by step: initial scoping/seed citation identification; keyword/phrase discovery; subject terms discovery; strategy logic; strategy evaluation; database search translation; grey literature search; and deduplication. **Figure 1** illustrates the SR search process with strategy development steps shaded boxes, and arrows indicating the iterative nature of steps.

**Figure 1.**
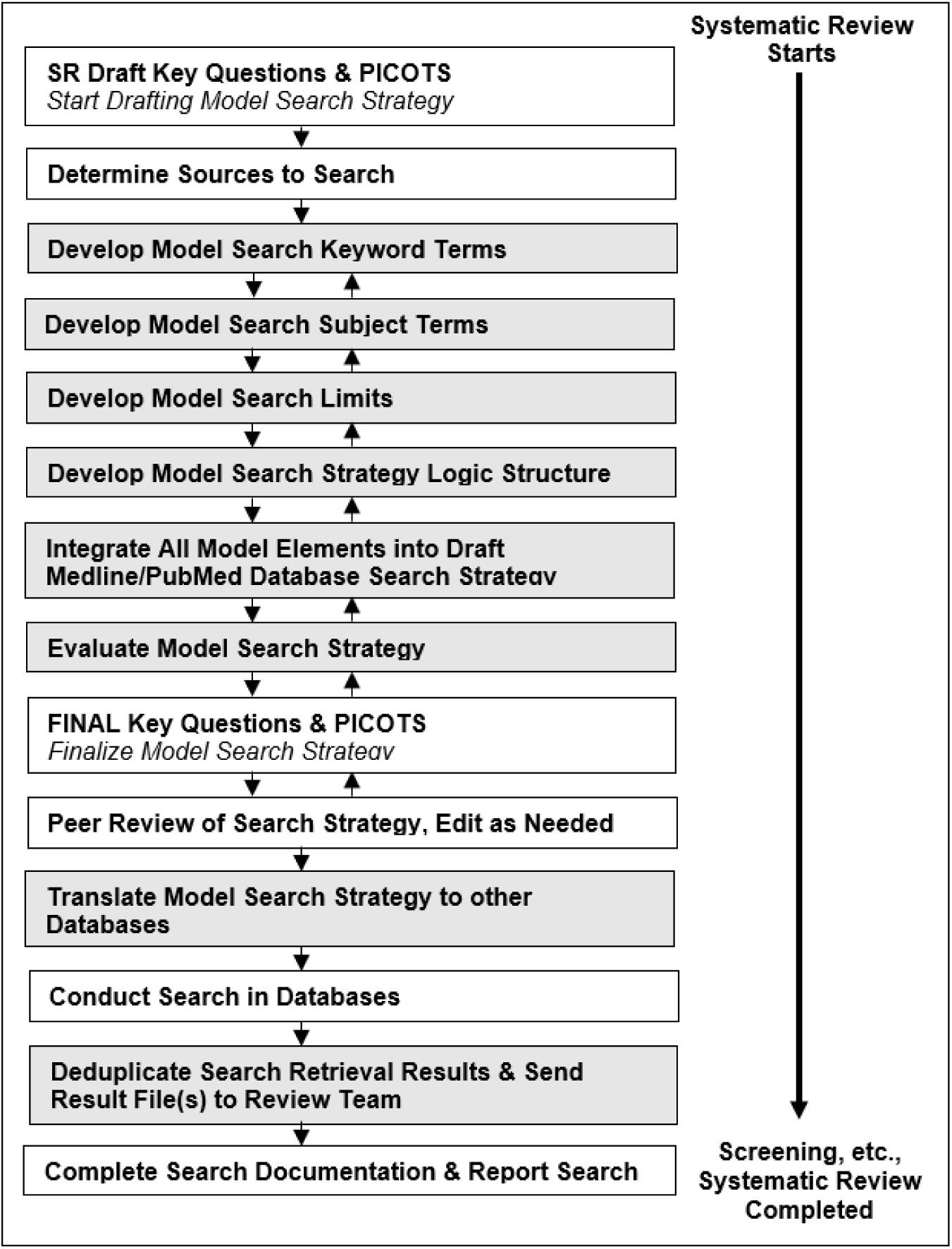
EPC Librarians’ Systematic Review Search Steps.

Lastly, we created a review form with the following domains for assessment: ease of use, access, user support, import parameters, and export parameters. These domains included objective and subjective criteria along with a narrative section providing notes on using the tool (please see **Appendix A. Review for Each Search Strategy Development Tool**).

### 2.1. Identification of Search Strategy Development Tools

In two searches of the SR Toolbox search software webpage (initial search review period September-November 2021 N=78; update search review period February-March 2023 N=24 new tools), we identified 102 search process tools.

### 2.2. Screening

Each team member independently reviewed the 2021 initial list of searching tools (N=78), suggested SSDT inclusion and exclusion criteria, and preliminarily assigned tools to a search strategy development step. As a team, we established final inclusion/exclusion criteria, then made consensus inclusion/exclusion and category assignment decisions for each tool. Two team members were assigned to review all tools within each search strategy step category, resolving any assessment disagreements via consensus. Category teams then presented their findings to the entire expert searcher panel for final review.

Each team member independently reviewed the 2023 list of new searching tools (N=24), then as a team we made consensus inclusion/exclusion decisions and categorized included tools.

#### 2.2A. Inclusion criteria

The tool supports one or more of the following search strategy development steps: initial scoping, keyword/phrase terms, subject terms, strategy logic, strategy evaluation, database translation, grey literature, and deduplication. A tool must also meet all of these additional criteria:

- Biomedical orientation
- Stand-alone tool (i.e., not integrated into a larger SR platform).
- Any method type (e.g., text-mining, machine learning, as well as ‘manual’ tools like Yale MeSH Analyzer)
- Created or updated within the last two years
- Freely available

#### 2.2.B Exclusion criteria

If a tool meets any one of the following criteria:

- Bibliographic (e.g., TRIP medical database), citation (e.g., Google Scholar), numerical, or statistical databases
- Citation management software (e.g., EndNote, Mendeley, or RefWorks)
- Fee or subscription-based tool
- Integrated tool within a SR platform (e.g., EPPI-Reviewer)
- Last updated ≥ two years ago
- No longer available
- Not a biomedical resource
- Not a search tool
- Not available during our review period (third quarter 2021 and first quarter 2023)
- SR screening or data abstraction tool (e.g., Distiller)
- Programming knowledge required to use it

### 2.4. Evaluation Form

Our evaluation form included both objective (e.g., tutorial availability) and subjective (e.g., tutorial effectiveness) measures. We developed the form based on our own experiences using the tools. A narrative review section includes discussion of the following:

- Bibliographic record fields for analysis (e.g., the entire record, title, subject terms)
- Methods to analyze the records (e.g., Boolean/truncation support, whether the search requires simplification, or the searching of individual fields only)
- Ease of translating the tool results back to the search (e.g., easy or difficult)
- Free-text general comments narrative section

### 2.5. Evaluations

All expert reviews for a given tool were combined for the overall score (see Results section, **Tables 1-5**). The overall score indicates whether a tool met all/most domain criteria, met half of the domain criteria, met less than half of the domain criteria, or was not available at the time of review/did not meet any domain criteria.

The evaluation form captured the time required to learn how to use the tool, along with the presence/absence of key features, including: access to the tool (web/download); user support (tutorial/help availability); ability to import (file/manual); and export formats. Within the user support section reviewers rated the usefulness of the tutorial/help features., e.g., if help was available but not very useful, the tool would be considered to meet less than half of that domain’s criteria.

In the narrative portion of the evaluation form, reviewers recorded more detailed notes on using each tool. We added an open-ended “general comments” section to capture reviewer impressions along with prompts to generate more focused comments on important aspects of tool functionality:

- What to analyze: Entire record | Title Field | Abstract Field | Subject Field
- Methods to analyze: Boolean/Truncation support | Requires simplifying search | Search fields individually
- Translation of Results Back to Search: Easy (result list cut & paste) | Difficult (how to interpret results unclear)

We later streamlined tool reviews as follows (NB: not all tools were assessed on each of these categories, as not all were applicable to every tool):

- WHAT: functions performed by the tool
- INPUT: file type(s)/other input(s) needed to import as the unit of analysis for the tool
- OUTPUT: file type(s)/other output(s) the tool exports
- HOW TO: tips and tricks on using the tool. If tool useful for multiple strategy development steps, advice for each type of usage. Work arounds for non-PubMed searchers as needed
- COMMENTS: reviewer impressions of its usefulness, usability, or problematic issues

## 3. RESULTS

Of the 102 search tools screened, 21 SSDT met our inclusion criteria (see **Figure 2** below). These 21 tools were grouped into search strategy development step categories: initial scoping/seed citation identification (7 tools), keyword/phrase (5 tools), subject term (4 tools), strategy logic (3 tools), strategy evaluation (3 tools), database translation (3 tools), grey literature search (1 tool), and deduplication (1 tool). Individual tools that can be used in more than one step appear multiple times. Please note: all the relevant SSDT within <u>Systematic Review Accelerator</u> platform were counted as individual tools). For the complete list of 102 tools please see Appendix B: Included and Excluded Search Tools. Figure 2. Flow Diagram

**Figure 2.**
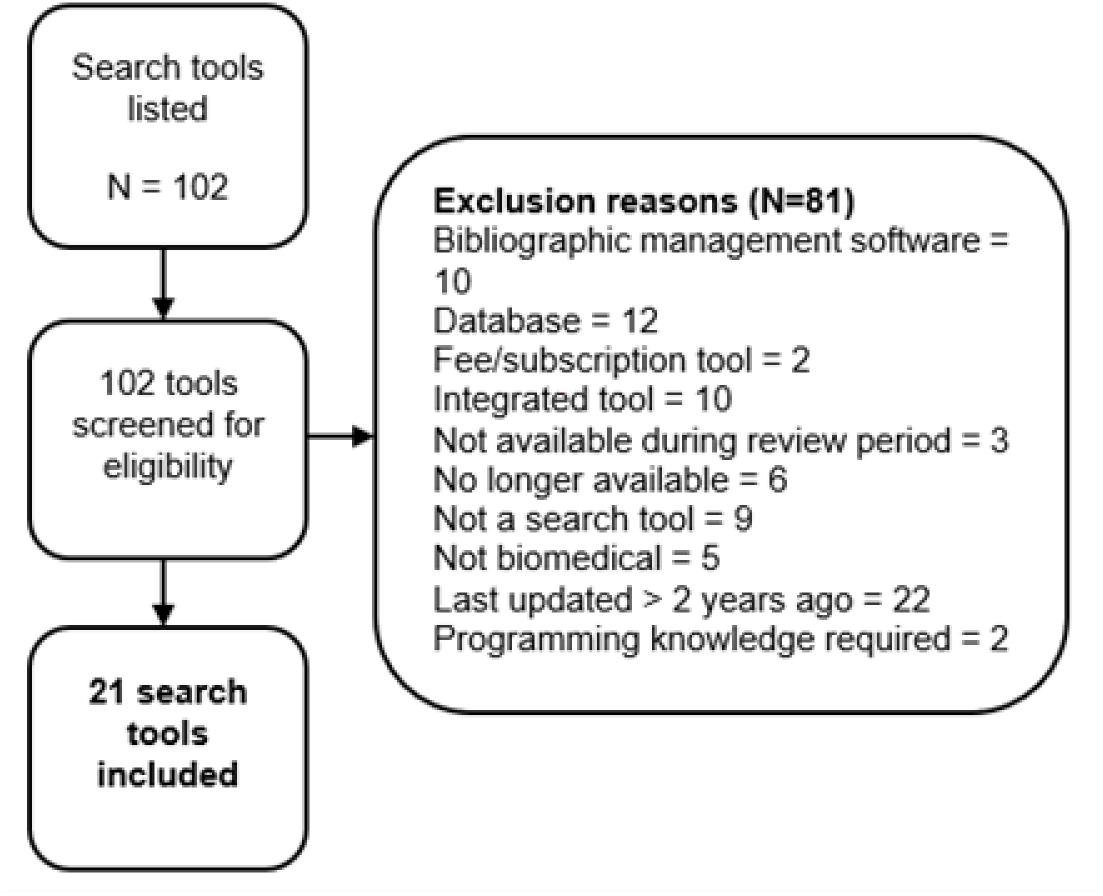
Flow Diagram.

**Tables 1-5** below show the summary ratings across all categories for each subgroup of tools. Assessments in these tables are expressed graphically, as follows:

**Table 1.**
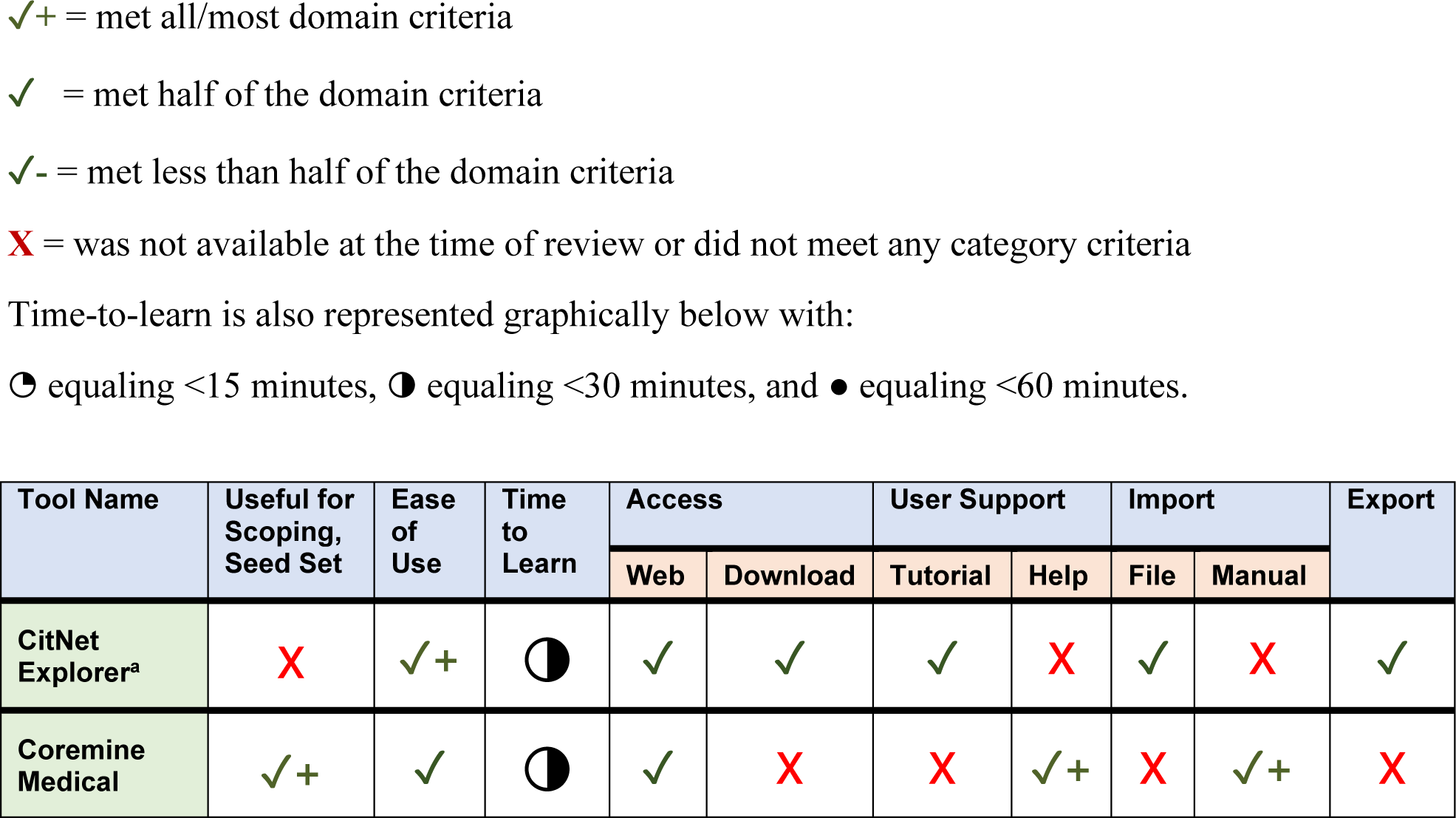
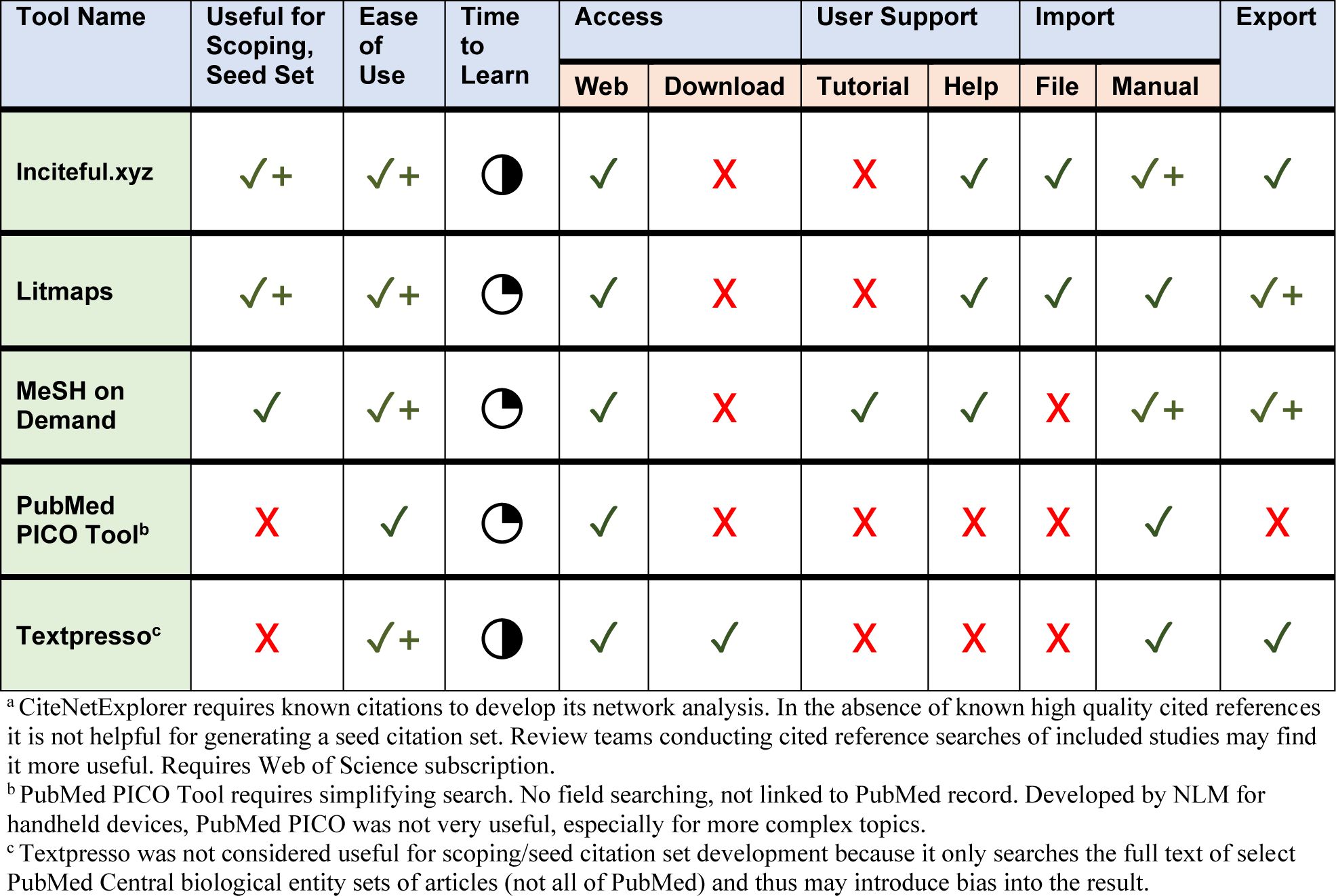

| Tool Name | Useful for Scoping, Seed Set | Ease of Use | Time to Learn | Access |  | User Support |  | Import |  | Export |
| --- | --- | --- | --- | --- | --- | --- | --- | --- | --- | --- |
|  |  |  |  | Web | Download | Tutorial | Help | File | Manual |  |
| CitNet Explorer<sup>a</sup> | X | ✓+ | ☉ | ✓ | ✓ | ✓ | X | ✓ | X | ✓ |
| Coremine Medical | ✓+ | ✓ | ☉ | ✓ | X | X | ✓+ | X | ✓+ | X |
|  |  |  |  | Web | Download | Tutorial | Help | File | Manual |  |
| Inciteful.xyz | ✓+ | ✓+ | ☾ | ✓ | ✗ | ✗ | ✓ | ✓ | ✓+ | ✓ |
| Litmaps | ✓+ | ✓+ | 🕒 | ✓ | ✗ | ✗ | ✓ | ✓ | ✓ | ✓+ |
| MeSH on Demand | ✓ | ✓+ | 🕒 | ✓ | ✗ | ✓ | ✓ | ✗ | ✓+ | ✓+ |
| PubMed PICO Tool<sup>b</sup> | ✗ | ✓ | 🕒 | ✓ | ✗ | ✗ | ✗ | ✗ | ✓ | ✗ |
| Textpresso<sup>c</sup> | ✗ | ✓+ | ☾ | ✓ | ✓ | ✗ | ✗ | ✗ | ✓ | ✓ |

<sup>a</sup> CiteNetExplorer requires known citations to develop its network analysis. In the absence of known high quality cited references it is not helpful for generating a seed citation set. Review teams conducting cited reference searches of included studies may find it more useful. Requires Web of Science subscription.
<sup>b</sup> PubMed PICO Tool requires simplifying search. No field searching, not linked to PubMed record. Developed by NLM for handheld devices, PubMed PICO was not very useful, especially for more complex topics.
<sup>c</sup> Textpresso was not considered useful for scoping/seed citation set development because it only searches the full text of select PubMed Central biological entity sets of articles (not all of PubMed) and thus may introduce bias into the result.

**Table 2.**
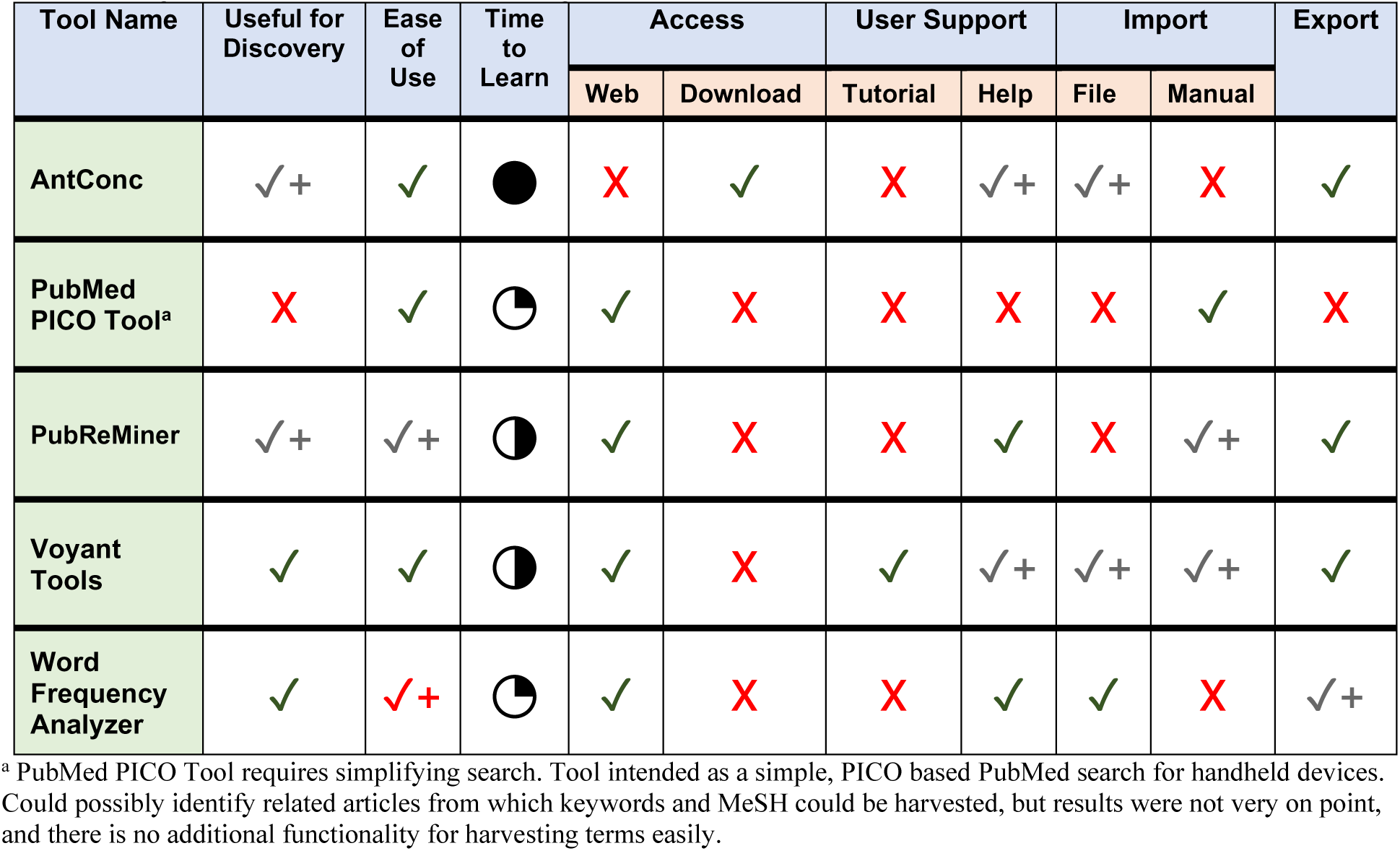
Keyword term/phrase discovery tools.

**Table 3.**
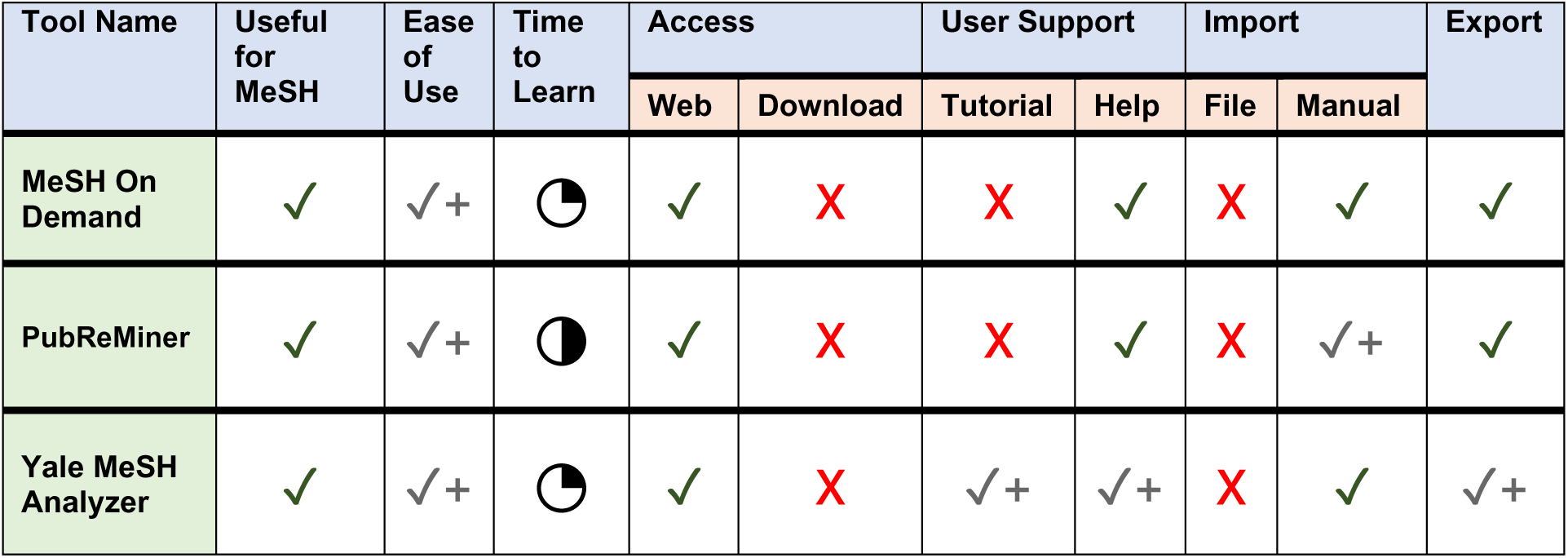
MeSH subject term discovery tools.

**Table 4.**
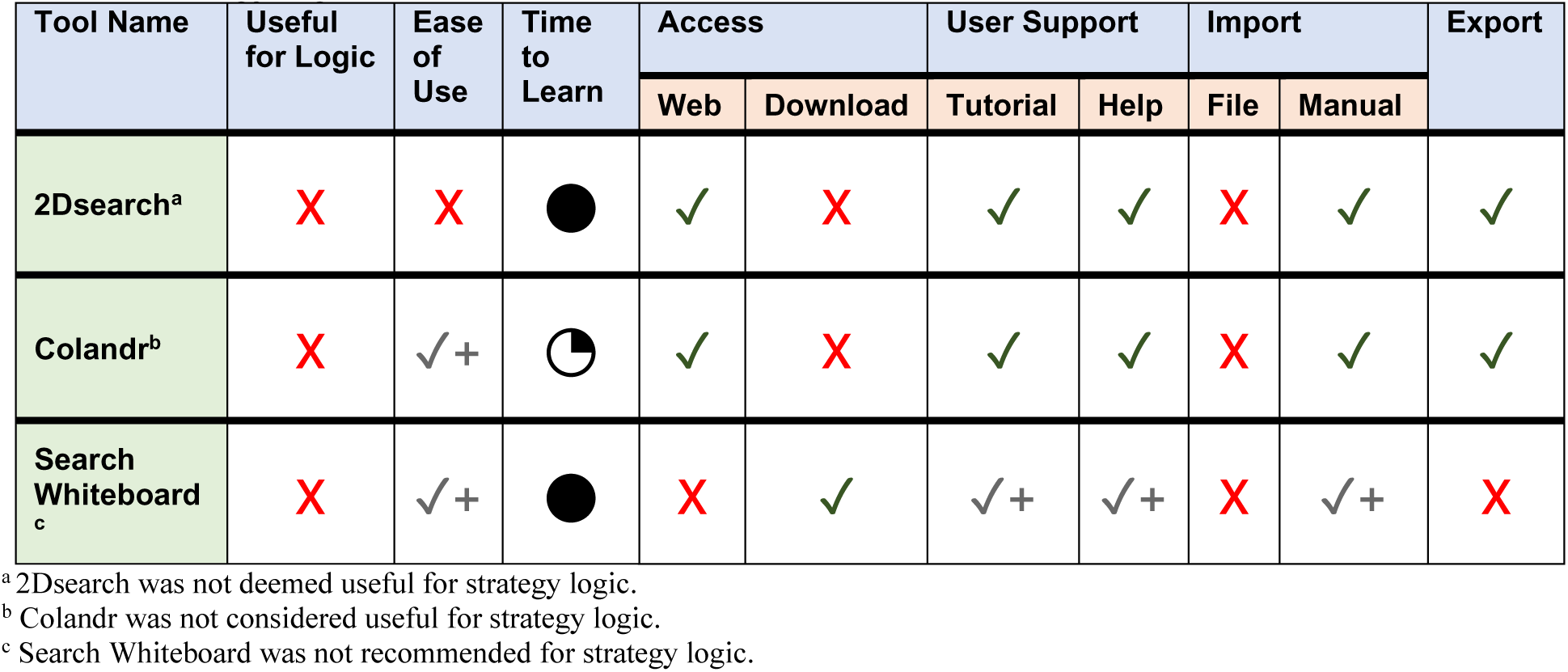
Strategy logic tools.

**Table 5.**
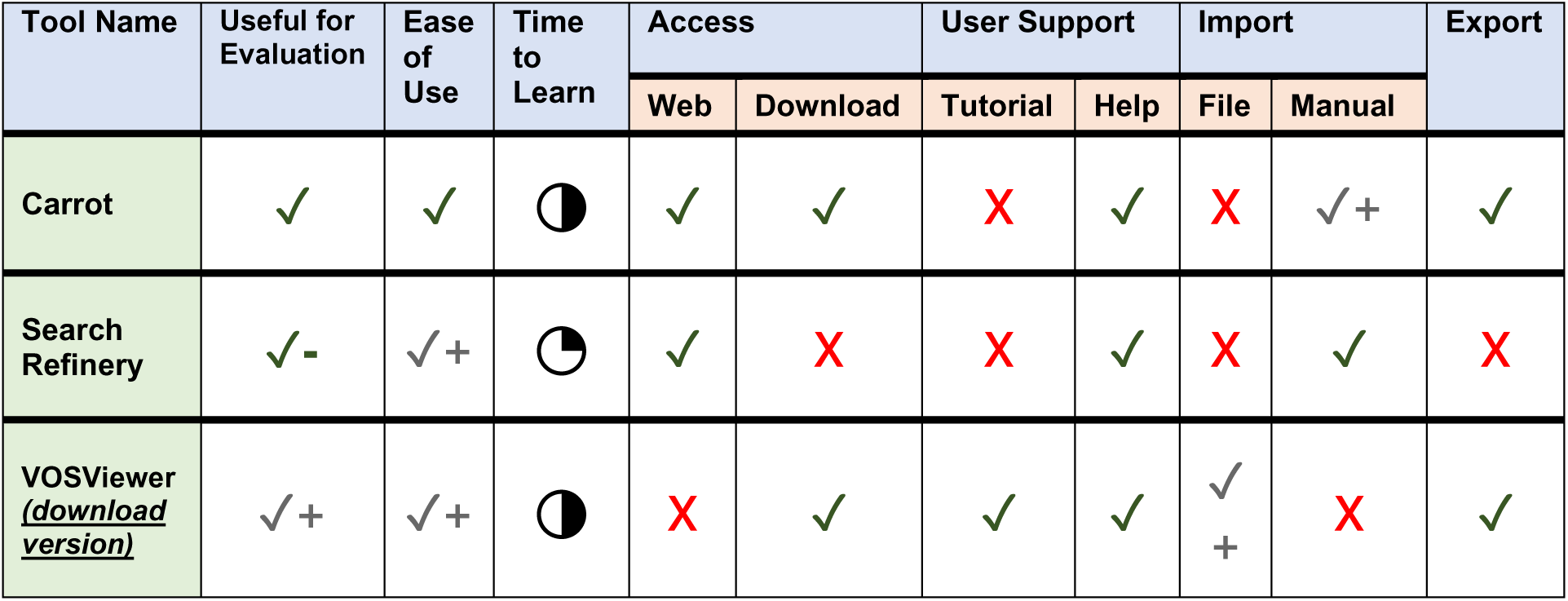
Strategy evaluation tools.

**Table 6.**
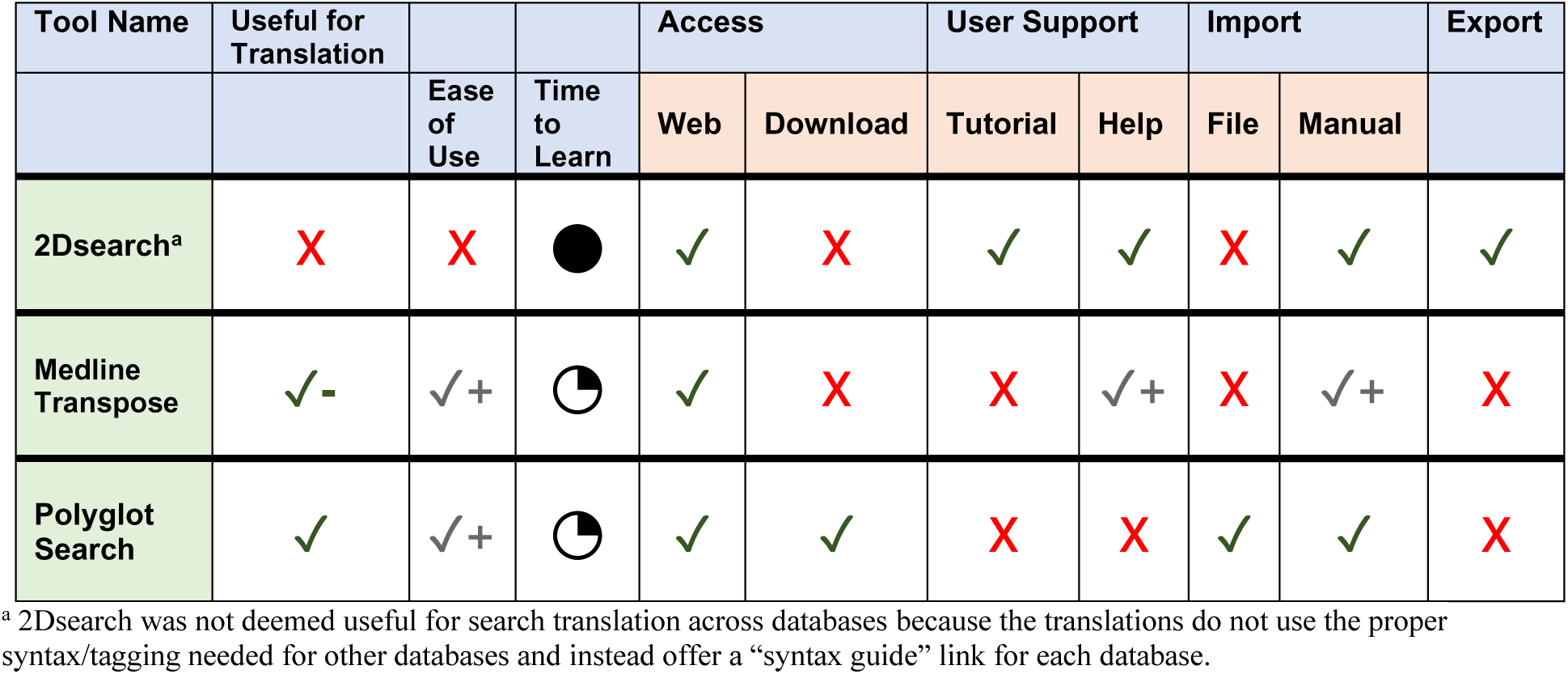
Search translation across databases tools.

**Table 7.** Grey literature search tool.

| Tool Name | Useful for Grey Literature | Ease of Use | Time to Learn | Access |  | User Support |  | Import |  | Export |
| --- | --- | --- | --- | --- | --- | --- | --- | --- | --- | --- |
|  |  |  |  | Web | Download | Tutorial | Help | File | Manual |  |
| 2Dsearch | ✓ | X | ● | ✓ | X | ✓ | ✓ | X | ✓ | ✓ |

**Table 8.** Search result deduplication tool.

| Tool Name | Useful for Deduping | Ease of Use | Time to Learn | Access |  | User Support |  | Import |  | Export |
| --- | --- | --- | --- | --- | --- | --- | --- | --- | --- | --- |
|  |  |  |  | Web | Download | Tutorial | Help | File | Manual |  |
| Deduplicator | ✓+ | ✓+ | ◐ | ✓ | ✓ | X | ✓+ | ✓+ | X | ✓+ |

In **Figure 3**, we provide an example of a narrative review.

**Figure 3.**
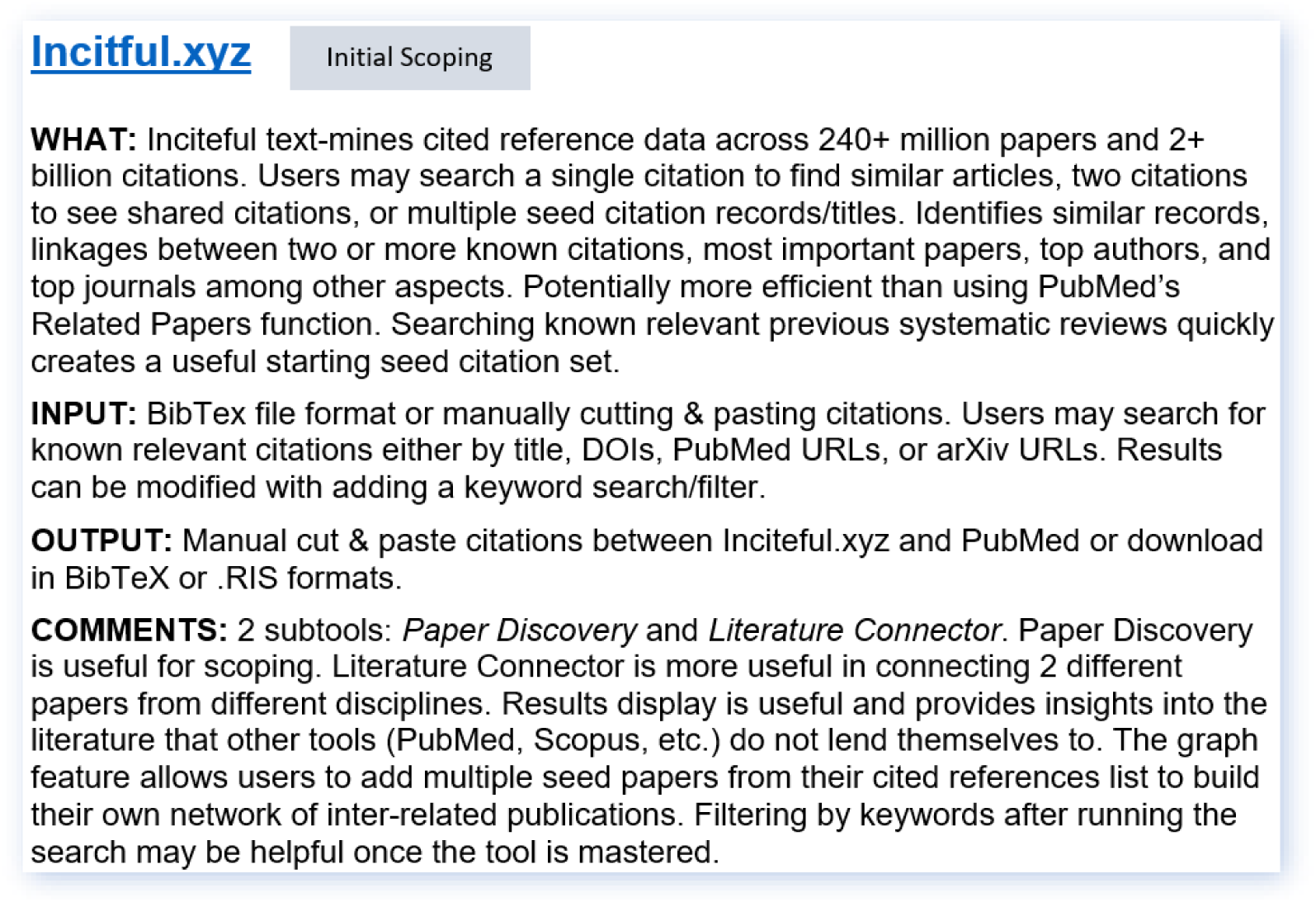
Sample Narrative Review.

In each case, we have attempted to provide sufficient information for an expert searcher to decide which tool(s) to use by search strategy development stage and understand tool requirements, capabilities, and limitations.

## 4. DISCUSSION

The use of search tools may increase efficiency, improve precision/sensitivity, and enable more timely delivery of results to review teams; though it should be noted that many of the 21 reviewed tools require workarounds to get usable results to inform strategy development. In four of the eight search strategy steps, at least one tool met all or most of the domain criteria. Notably, in the search strategy logic category (**Table 4**) all three tools did not meet any of the category criteria, i.e., assisting searchers to create or test their search logic. **Tables 1-3** include the three initial search development steps, which are also the categories with the largest number of available, highly rated tools. These steps lend themselves to tools that analyze structured bibliographic records using word frequency and cited reference linking. Historically, searchers reviewed records “manually” to identify keywords, check cited references, etc.; these tools make it possible to perform these tasks more quickly across a larger sample of records, thereby improving the timeliness and comprehensiveness of the search. Four of the seven tools evaluated for initial scoping/seed citation set development (**Table 1**) were considered useful, while three were not. We especially favored Inciteful.xyz for quickly capturing relevant SR included study citations to use as a seed set to test strategy retrieval. Among the keyword/phrase tools PubReMiner is a quick and easy way to identify common keywords in title/abstract and MeSH fields respectively, while AntConc is especially useful for finding phrases and setting adjacency limits. Strategy logic (**Table 2**) included three tools, none of were able to test strategy logic for either simple or complex searches. Each of the MeSH discovery tools (**Table 3**) performed well as designed. For searchers wanting to identify common MeSH terms across a large sample of relevant PubMed records (up to 10,000) PubReminer is the preferred solution. MeSH on Demand supplies likely MeSH terms based on a small amount of text (say an abstract) and Yale MeSH Analyzer displays the MeSH terms for only 20 PMIDs at a time. The search strategy evaluation (**Table 4**) and deduplication (**Table 5**) steps also include a number of useful tools that analyze structured bibliographic records, via results clustering and matching respectively. The strategy evaluation tools identify relevant and irrelevant clusters in retrieved sets. Retrieved irrelevant records can be analyzed to determine whether it is possible to ‘NOT’ them in of the search strategy, thereby decreasing the review team’s screening burden. While VOSviewer took longer to learn than some others, it ranked the highest overall across all domains. Tools in the search translation across databases category (**Table 5**) were found to be primarily useful for translating search string syntax (the relatively easy piece to translate). For searchers using common database platforms, e.g., Ovid, EBSCOhost, Embase, etc., this can save time over manual editing. Please note: searchers must know their selected databases’ syntax to troubleshoot errors. Also, these tools do not translate subject terms between databases (e.g., MeSH terms into CINAHL subject terms). Lastly, grey literature searching (**Table 5**) includes only one tool, 2Dsearch, which saves search strings for multiple grey literature websites. This can save time for searchers who regularly search particular grey sources.

We used sample simple and complex SR topic search strategies to determine how well the tools perform under varying conditions. Of the 21 reviewed tools most required searches to be broken down into individual components.

## 5. LIMITATIONS

The primary limitation of this evaluation is that we selected tools to review from those available on SR Toolbox. While we consider the SR Toolbox catalog to be fairly comprehensive, we did not search for tools elsewhere and therefore may have missed some potential candidates. Further, we chose not to evaluate all the tools in the SR Toolbox that are categorized as search related.

These included: tools we deemed not relevant to the biomedical field; tools whose website was non-functional, that required programming knowledge to use, that required the download of software, or that were not permitted due to local security firewalls. We also chose not to evaluate tools that: 1) had not been updated within the last two years (instead focusing on tools with active support and feature enhancement); 2) fee-based tools; and 3) tools that we believe were out of scope (e.g., screening tools).

A secondary potential limitation was the varying levels of prior experience with some of the tools among our team of investigators. Working knowledge (or lack thereof) may have resulted in different learning curves, and thus different ratings for some of the tools. Further, the general experience level of our team could mask issues a relative newcomer to systematic searching may find confounding or prohibitive.

Since the first quarter 2023 search and review period there has been rapid development of generative artificial intelligence tools for SRs. This latest category of search tools is not included in this guide.

Finally, creating an evaluation form to use across all categories of tools required discussion and compromise. As a result, we took a more general approach to evaluating all tools with the same criteria. This created some challenges in terms of how to note or score unique features within some of the tools. Some tools may have more utility than our review captures.

## 6. FUTURE RESEARCH

Future review of these tools may benefit from updated evaluation protocols for each specific category of tools. Focusing evaluations on the unique aspects of the various categories (i.e., scoping versus database translation) would allow for a more detailed review of the specific functionality of tools. Our expert searchers agreed that it is critical to consider the local work environment while contemplating which tool(s) to integrate into a searcher’s workflow. Regular use of SSDT may help searchers develop confidence and trust in them. Further research into whether their consistent use results in shorter turn-around times for literature searches is needed. Further evaluation of which search platform, e.g., OVID Medline instead of PubMed, is more conducive to using SSDT is needed. Lastly, more formal investigation of best practices for integrating SSDT into the SR workflow is needed.

In addition, our analyses indicate that there is a need for the development of tools to assist in all steps of SR search creation, in particular there are no or very few good tools for evaluating strategy logic, translation of search syntax/subject terms across databases, and developing searches for grey literature websites.

## 7. CONCLUSION

Software tools have the potential to improve the search-design process for evidence synthesis products (SRs, rapid reviews, evidence maps, etc.). We present a set of graphical evaluation tables of the 21 tools to help searchers quickly identify the tools available to them at each step of the search strategy development process and to connect to their preferred tool(s). We supplement the graphical tables with more detailed narrative reviews describing how to use the tools.

## Data Availability

All data produced in the present work are contained in the manuscript

## Acknowledgements

The authors gratefully acknowledge the following for their contributions to this project: Bendte Fagge, Scott Macdonald, and Ed Reid.

## Funding Disclaimer

This manuscript is based on research conducted by the Scientific Resource Center under contract to the Agency for Healthcare Research and Quality (AHRQ), Rockville, MD (Contract No. HHSA 290-2017-00003C, 75Q80122C00002) and the following Evidence-based Practice Centers: Brown University (Contract No. 75Q80120D00001/75Q80120F32003), ECRI (Contract No. 75Q80120D00002), Johns Hopkins (Contract No. 75Q80120D00003), and RTI-UNC (Contract No. 75Q80120D00007). The findings and conclusions in this document are those of the authors, who are responsible for its contents; the findings and conclusions do not necessarily represent the views of AHRQ. Therefore, no statement in this report should be construed as an official position of AHRQ or of the U.S. Department of Health and Human Services.

The authors have no competing interests to declare.

## Appendix A. Review for Each Search Strategy Development Tool

### 2Dsearch [DATABASE TRANSLATION, GREY LITERATURE, STRATEGY LOGIC]

**WHAT:** Visual “search canvas” approach to developing search strategies, rather than command line. The tool automatically searches PubMed, but other databases/search engines are available to select (including Google Scholar) or users can add preferred sites, especially useful for grey literature sites.

**INPUT:** Options to enter a search into 2Dsearch (from the menu):

a. Save As: saves search from the canvas and then the search is saved for later use (using the Open command to rerun the search)
b. Enter Query String: Paste a search to run it. This adds the search in canvas format and it can be saved rerun as described in a) above.
c. Create the search on the canvas de novo.

**OUTPUT:** Search result list cut & paste. Save search strategies, share search string URLs, copy query strings for reporting purposes, and customize frequently used grey literature sites. Search translation is easy. Search strings can be saved or cut and pasted into search reporting document. Searches are automatically conducted in target database(s) or website(s), so after creating the search users can view/download results there. If using multiple sites having automatic syntax translation can save time and searcher frustration over forgotten syntax rules. Searchers can add/save websites (save query strings and/or grey literature websites for quick searching in future).

**HOW TO: GREY LITERATURE METHODS:** Existing 2Dsearch strategies must be parsed to fit within grey literature site searching constraints. Syntax guides are available for each linked source. Preselected popular grey literature sites include Epistemonikos and TRIP Database. Especially useful for routinely searched grey literature websites.

**HOW TO: STRATEGY LOGIC METHODS:** Import existing PubMed search strategies allows checking for search errors, especially duplication of terms or MeSH terms.

**COMMENTS:** Experienced searchers used to conventional command line searching may find the ‘search canvas’ approach more time consuming to develop a search. Users creating searches/adding sites for the first time will likely encounter issues requiring trial and error to resolve, but thereafter they can be saved for future use. Visually-oriented searchers might find the canvas approach refreshing.

### AntConc [KEYWORD/PHRASE]

**WHAT:** Originally a linguistic studies concordance tool, AntConc has a suite of eight user controlled textual analysis tools. Especially useful for diffuse/broad review topics with many potential keywords/phrases, or when there are a large number of known relevant citations that would take too much time to review individually for useful search terms.

**INPUT:** Text file (also known as a corpus) of identified highly relevant database records (more records better than fewer).

**OUTPUT:** Search translation results are easy. Export results as text or data tables. Alternatively, note useful terms, phrases, and especially adjacency numbers.

**HOW TO: KEYWORD DISCOVERY:** Save title and abstract fields as separate text files because AntConc parses everything together, thus losing the ability to discriminate results by field. You may want to edit out noise words in notepad (e.g., “title,” “abstract” if added to each record by database as well as “the,” “and,” etc.), before importing into AntConc. After importing a file (a.k.a., creating a quick corpus), click on the *Word* tab and then the start button to see the terms list ordered by frequency.

**HOW TO: PHRASE DISCOVERY:** Import abstract or combined title/abstract text files. The *Cluster and N-grams* tabs are useful to identifying phrases and establishing optimal term adjacency numbers.

**COMMENTS:** Offering excellent performance, AntConc has a steep learning curve and is difficult for those unfamiliar with linguistic terms to use (especially if used infrequently).

### Carrot^2^ [STRATEGY EVALUATION]

**WHAT:** Text-mining tool whose algorithm visually clusters PubMed records into topic groupings. Identifying irrelevant topic clusters that can be excluded (‘NOT’ed out) from strategy to improve search precision.

**INPUT:** Analyzes a PubMed search strategy. Requires users to simplify the search to one or two PICO elements. PubMed syntax (i.e., AND, [TIAB]) is supported and may be used to focus results.

**OUTPUT:** Search translation is moderate to difficult. Some manipulation of results is required. The tree-map and pie-chart visualizations provide novel clustering views of research which may display connections of unrelated concepts to the review topic (e.g., broad or widely used terms).

**COMMENTS:** First-time and infrequent users may have difficulty interpreting the search output. Uncertainty around whether pared down search adequately clusters results to discern unrelated concepts.

### CitNetExplorer [INITIAL SCOPING]

**WHAT:** Visualization of citation networks

**INPUT:** Web of Science .txt or tab-delimited files

**OUTPUT:** Citation network can be exported in Pajek file format

**HOW TO:** Initial visualization of citations can be further analyzed by 1) drilling down/expanding based on marked citations to create a subnetwork; 2) clustering publications based on their citation relations; 3) core publications defined as publications with X number of citation relations with other core publications.

**COMMENTS:**

- Easy to translate results back to search
- More useful after main search, when cited reference searching included studies.
- Must install JAVA to run the download or access the web version.
- Other tools like Inciteful.xyz are easier to use and do not require subscriptions to Web of Science

### Colandr [STRATEGY LOGIC]

**WHAT:** Searchers add terms into their template which then creates the Boolean structure.

*Please note: it puts all terms within quotes*.

**INPUT:** Input keywords and Colandr tries to create a Boolean search string.

**OUTPUT:** Tool generated Boolean search strings connected only with the “OR” operator.

**COMMENTS:** The Boolean logic was incorrect and the tool functionality does not provide the search in platform specific syntax. As a result, a great deal of editing is required for each strategy. A Gmail account is required to access the help and tutorial files.

### Coremine Medical [STRATEGY EVALUATION]

**WHAT:** Users enter search terms which are then mapped to Coremine Medical’s ontology. While not useful for analyzing an existing search per se, it does provide some context of drugs, topics, etc. connected to any group of topics. May provide the user with more “AND” combination suggestions.

**INPUT:** Strategies require simplification to a single word, phrase, or MeSH term. There is no field searching, however, Boolean (AND, OR) is supported. Users may combine topics iteratively, but it does not support complex search strings.

**OUTPUT:** Translation of results back to the search is moderate to difficult. With this tool, the user is not really evaluating the performance of the search, but rather an overly simplistic version of the search. Some manipulation of the results is required.

**COMMENTS:** Tool is useful for searching for author names and citations. It is perhaps most useful for citation-type analysis and compiling a list of authors writing in a particular field.

### Deduplicator (Systematic Review Accelerator) [DEDUPLICATION]

**WHAT:** Deduplicates based on record fields that include DOI and PMID.

**INPUT:** Requires an EndNote import file. Three settings for review for duplicates: cautious, balanced, or thorough.

**OUTPUT:** Users export now deduplicated file to upload to EndNote.

**COMMENTS:** Worked very quickly on the test file, no error messages. Requires extensive manual review of automated matching.

### Incitful.xyz [INITIAL SCOPING]

**WHAT:** Inciteful text-mines cited reference data across 240+ million papers and 2+ billion citations. Users may search a single citation to find similar articles, two citations to see shared citations, or multiple seed citation records/titles. Identifies similar records, linkages between two or more known citations, most important papers, top authors, and top journals among other aspects. Potentially more efficient than using PubMed’s Related Papers function. Searching known relevant previous systematic reviews quickly creates a useful starting seed citation set.

**INPUT:** BibTex file format or manually cutting & pasting citations. Users may search for known relevant citations either by title, DOIs, PubMed URLs, or arXiv URLs. Results can be modified with adding a keyword search/filter.

**OUTPUT:** Manual cut & paste citations between Inciteful.xyz and PubMed or download in BibTeX or .RIS formats.

**COMMENTS:** 2 subtools: *Paper Discovery* and *Literature Connector*. Paper Discovery is useful for scoping. Literature Connector is more useful in connecting 2 different papers from different disciplines. Results display is useful and provides insights into the literature to which other tools (PubMed, Scopus, etc.) do not lend themselves. The graph feature allows users to add multiple seed papers from their cited references list to build their own network of inter-related publications. Filtering by keywords after running the search may be helpful once the tool is mastered.

### Litmaps [INITIAL SCOPING]

**WHAT:** It may be useful for developing a scoping search or identifying citations, though PubMed and Scopus/Web of Science are better for this purpose. It may be useful in search generation (through citation searching) but not helpful for refining topics.

**INPUT:** Requires dramatically simplifying search strategy.

**OUTPUT:** Translation of search results is difficult. How to interpret results is unclear.

**COMMENTS:** The tool is flexible and powerful for what it does. No information is provided on sources searched.

### Medline Transpose [DATABASE TRANSLATION]

**WHAT:** Converts search strategy syntax between Ovid MEDLINE and PubMed. Does not convert into any other bibliographic database syntaxes. Ovid searchers can more efficiently convert their search to PubMed syntax for use with other search strategy development text-mining tools.

**INPUT:** Cut & paste entire Ovid MEDLINE (or PubMed) search strategy as written.

**OUTPUT:** The documentation tab outlines how Medline Transpose interprets syntax and any known issues translation issues by specific field. Javascript converts from one syntax to another and inserts Boolean connectors in at the end of analysis.

**COMMENTS:** Expert searchers should be aware of possible errors in the search translation and review the translated search strategy for accuracy. Not recommended for inexperienced searchers with limited understanding of PubMed and Ovid search syntax. Following the translated strategy are notes explaining problems with translation (Ovid .ti,ab searches fewer fields than PubMed [tiab] for example). Best used with Firefox, Chrome, or Safari browsers.

The “Documentation” section serves as a help function. It is very clearly written but could benefit from some example results and/or a tutorial. There are many errors for complex searches which requires searchers to review results closely. The table converting tags between PubMed and OVID Medline is useful. Translating from OVID to PubMed may be more problematic than translating PubMed to OVID.

### MeSH on Demand [INITIAL SCOPING, SUBJECT TERM]

**WHAT:** Maps submitted text to suggested MeSH terms.

**INPUT:** Cut & paste any English language text with topical content (abstract, protocol, etc.). Limit: 10,000 characters. Easy to use and understand, however submitting representative, descriptive text is critical to achieving best results from the tool.

**OUTPUT:** Easy to cut and paste, save results as a .txt file, or click hyperlinks for MeSH and PMID links.

*Please note: exported result text file do not contain hyperlinks to MeSH or similar article PMIDs*.

**HOW TO: INITIAL SCOPING** If submitted abstract retrieves too many MeSH terms, try searching the objectives/background separately to other sections.

Suggested MeSH terms appear next to your entered text while similar articles are listed below. Review and note the useful terms from the presented list (terms listed in ranked weighted order) and/or view suggested related articles. ‘Start PubMed Search’ button List of all identified MeSH terms (population, intervention, etc.) is generated by selecting the ‘Start PubMed Search’ button (terms are subsequently searched in PubMed). This may not a useful option since terms are automatically combined with “AND” and there is no option for other combinations. It is useful to note terms of interest or use the ‘Export Data’ button to record suggested terms and similar articles for later use.

**HOW TO: SUBJECT TERM DISCOVERY** As noted above, submitting representative, descriptive text is critical to achieving best results from the tool—as well-crafted review protocol text is typically unavailable when the search is being created expert searchers will very likely prefer to use PubMed’s MeSH database to develop specifically tailored MeSH terms for their topic.

**COMMENTS:** The suggested MeSH terms rely on the accuracy of the submitted review topic text. In our testing, the MeSH terms are more accurate and complete for simple topics than for complex ones. Suggest similar articles are limited to 10. Processing of the search depends on the length of the submitted text, for example a 350-word abstract took 2 minutes when we tested.

### Polyglot Search (Systematic Review Accelerator) [DATABASE TRANSLATION]

**WHAT:** Translates database-specific search strategy syntax between databases, e.g., input an Ovid Medline search for translation into EBSCOhost CINAHL syntax.

**INPUT:** Text of PubMed or OVID Medline search strategy. Paste in text to be analyzed or upload from a .txt file.

**OUTPUT:** Cut & paste the search strategy, however searchers must review the search for errors. There are parts of a strategy that the tool is not able to translate, like field tags. Most notably are line logic errors and translation of MeSH terms to non-MeSH databases.

**COMMENTS:** Expert Searchers should review the translated search strategy and syntax for accuracy. Not recommended for inexperienced searchers with limited understanding of search syntax in target databases. Be aware that the Polyglot Search translates search syntax between databases but does not adapt subject terms across databases, i.e., MeSH terms are translated into Ovid APA PsycInfo syntax erroneously and there is no error message to inform the user.

Translation between the following databases is available: PubMed, Ovid MEDLINE, Cochrane Library, Embase (Elsevier, Ovid), CINAHL (EBSCOhost), Web of Science (basic and advanced search), Scopus (basic and advanced search), APA PsycInfo (Ovid), ProQuest Health and Medical, SPORTDiscus, lexical Tree JSON. The Help section has tutorial information and screenshots but does not cover how to import from a .txt file and there does seem to be a file size limit. The checkbox to replace line references does not always work. Difficult to find error “bubbles” because the dark blue font used to indicate them is also used for keywords. Works better for simpler searches topics.

### PubMed PICO Tool [INITIAL SCOPING, KEYWORD/PHRASE]

**WHAT:** A simple, PICO based search of PubMed for handheld devices.

**INPUT:** Keywords or terms are inserted into PICO fields. The tool does not accept phrase searches with quotation marks. “Ask Medline” (help function) is available if no results are identified.

Requires simplifying the search to single keyword or short phrase. No Boolean, truncation, or field searching capabilities. The search requires simple text words search but users may use multiple text words at a time.

**OUTPUT:** Results are summaries of PubMed records. There is no option to download citations. User must cut & paste results into a document. Table, abstract, publisher full-text, and related links are provided.

It is difficult to utilize results of the search. Results are citations and appear in a list. No data on interpretation of search terms is available.

*Please note: does not link to PubMed record*.

**COMMENTS:** Developed by NLM for handheld devices in clinical situations, PubMed PICO does not readily translate to systematic review (SR) scoping searches or development of seed citation sets, especially for more complex topics. No added value for experienced PubMed searchers. The lack of help, instructions, and export features are hindrances. Results are unpredictable depending on specificity of the terms used (Boolean AND helps with producing useable results). Works better for simple topics. PubMed would be preferred for scoping work related to systematic reviews or anything even moderately complex.

Could possibly identify related articles from which keywords and MeSH could be harvested, but results are not on point, and there is no additional functionality for harvesting terms easily.

### PubReMiner [KEYWORD/PHRASE, SUBJECT TERMS]

**WHAT:** Word frequency analysis of input records allows for quick identification of high value keyword (title, title/abstract, etc.), MeSH terms, drug names, etc.

**INPUT:** Seed citation (known relevant) PMIDs or conduct a simplified PubMed syntax search.

**OUTPUT:** Click on linked MeSH term(s) to add to the search. PubMed search string may be edited as needed before utilizing the ‘Go to PubMed with Query’ button. On single terms of interest, users click on the blue ‘P’ to view PubMed MeSH search results.

### HOW TO: KEYWORD TERM DISCOVERY

<u>Method 1:</u> PubMed syntax searches are supported, including Boolean, truncation, and individual fields. Simple or complex search strings supported.

The tool builds a term frequency table based on the PubMed records. The tool lists results in various tables based on journal title, author name, and text terms.

<u>Method 2:</u> Paste PMIDs into the Enter Your PubMed Query box, click Start PubReMiner button. After initial results display users can select which columns to display on the right side of the screen; after selection click on Search with Manual Adjustment button. Recommended columns to display: substance, title, title/abstract, title/abstract/major heading/substance.

**HOW TO: PHRASE DISCOVERY** Not applicable for phrase discovery.

### HOW TO: SUBJECT TERM DISCOVERY

PubMed syntax searches are supported, including Boolean, truncation, and individual fields. The tool builds a term frequency table of MeSH terms from identified PubMed records. Simple keyword search strings may work best to aid MeSH term discovery. For subject term discovery both the MeSH and substance fields (depending on review topic) are the most useful.

This tool is simple to use for identifying and reviewing potential MeSH terms. Compared to other tools, this analyzes more records and its result display is easier to understand and manipulate.

Expert searchers will want to use PubMed’s own MeSH database to develop a comprehensive list of terms for their search strategy.

Ovid searchers may create PubMed search strings or export PMIDs list from your Ovid search to utilize the tool.

**COMMENTS:** While this tool does analyze the results of a search and provide counts of words and phrases found in titles and abstracts, the usability of the information for refining searches is limited.

Instructions help somewhat but the steps are not clearly defined. There is a tutorial on YouTube that provides a quick overview of how to best utilize the tool. May be marginally useful to an intermediate to experienced user. Some functionality is unclear, including (determining) which abstracts are used to generate tables and how Abstract Limit works. Results may be too long for complex topic to be useful. Page not updated since 2014. FAQ seems out of date.

### SearchRefinery (Systematic Review Accelerator) [STRATEGY EVALUATION]

**WHAT:** Generates a search strategy visualization, with bolder lines between terms representing a greater number of search results.

**INPUT:** Copy and paste a PubMed search strategy into the search query box, then add seed citation PMIDs (if desired). Requires simplifying a PubMed search by ‘AND’ing together multiple line searches in the tool, and spelling out words, i.e., not using truncation.

**OUTPUT:** The output is a visualization of the number of shared citations between concepts allowing searchers to see the number of results generated by each term combination.

**COMMENTS:** Conversion of Ovid Medline syntax to PubMed syntax may result in errors. This may provide a broad view of shared results (though you can do this in native platform as well). Free registration is available.

### Search Whiteboard [KEYWORD/PHRASE, STRATEGY EVALUATION]

**WHAT:** This is an Excel file. Searches are run in various user selected databases from within this downloaded Excel tool. Results are loaded into a new browser tab, organized by resource (PubMed, TRIP, etc.). This ensures that the same strategy is consistently used. Users can keep track of searches all in one place.

**INPUT:** User may add terms to columns by double clicking on the terms of interest or they may enter search terms into the search box. Using prespecified strategies from sources (i.e., covid strings) is also an option.

### HOW TO: KEYWORD TERM DISCOVERY

Difficult to determine the direct applicability to keyword term discovery. The simple search is effective and uncomplicated. The intermediate search has many more options to utilize and is more complicated to understand. It may be difficult for the user to determine how to effectively use the more complicated search function(s).

### HOW TO: PHRASE DISCOVERY

Difficult to determine the direct applicability to phrase discovery. The simple search is effective and uncomplicated. The intermediate search has many more options to utilize and is more complicated to understand. It may be difficult for the user to determine how to effectively use the more complicated search function(s).

### Textpresso [INITIAL SCOPING]

**WHAT:** Identify search terms in full text materials in select PubMed Central (PMC) Open Access subsets. Useful to identify PMC articles mentioning specific medical devices, procedures, tests, etc. that are not included in abbreviated PubMed/PMC records; however, the PMC search interface is more user friendly, offering more search, selection and download options.

**INPUT:** Simple keyword search within a sentence or across the entire document.

**OUTPUT:** Copy and paste desired citation PMIDs into PubMed.

**HOW TO:** Recommend sentence level searches and simplifying searches down to single concepts. Alternatively, search for PICO elements, e.g., smokers (population) AND lung cancer (indication). Search allows Boolean logic and case sensitive search options. Truncation and other sophisticated search refinements are available. Searching the full-text yields non-relevant results, even for a simple Boolean AND search. It is Interesting to see KWIC in the full-text but the Annotation and Curation features are difficult to understand. The tool uses Drupal/Lucene syntax.

Select one citation to view (or save) at a time, then click the curation tab to view keyword in context of your search terms. View individual full text materials, annotate them and later export to other bibliographic systems.

## Appendix B. Included and Excluded Search Tools

**Included Tools**

1. 2Dsearch
2. AntConc
3. Carrot^2^
4. CitNetExplorer
5. Colandr
6. Coremine Medical
7. Deduplicator (Systematic Review Accelerator)
8. Inciteful.xyz
9. Litmaps
10. Medline Transpose
11. MeSH on Demand
12. Polyglot Search (Systematic Review Accelerator)
13. PubMed PICO Tool
14. PubReMiner
15. Search Refinery (Systematic Review Accelerator)
16. Search Whiteboard [Excel file]
17. Textpresso
18. VOSviewer
19. Voyant Tools
20. Word Frequency Analyzer (Systematic Review Accelerator)
21. Yale MeSH Analyzer

**Excluded Tools**

1. **A2A (Apples to Apples)** exclusion reason: last updated >2 years ago.
2. **Aigaion** exclusion reason: bibliographic management software.
3. **AlvisIR Food Semantic Search Engine** exclusion reason: no longer available.
4. **Article-based PubMed Search Engine (APSE)** exclusion reason: no longer available.
5. **Article Galaxy** exclusion reason: it is a database, not a search strategy development tool.
6. **Author Name Disambiguation (AND)** exclusion reason: not a search tool.
7. **Automatic VAriant Evidence DAtabase (AVADA)** exclusion reason: no longer available.
8. **BADERI** exclusion reason: last updated >2 years ago.
9. **Bebop** exclusion reason: bibliographic management software.
10. **BEST (Biomedical Entity Search Tool)** exclusion reason: last updated >2 years ago.
11. **Buhos** exclusion reason: search tool part of integrated review platform.
12. **CADIMA** exclusion reason: search tool part of integrated review platform.
13. **Cientopolis Scolr** exclusion reason: search tool part of integrated review platform.
14. Citation and text-based framework for retrieving publications for literature reviews exclusion reason: last updated >2 years ago.
15. **Citationchaser** exclusion reason: it was not available during review period.
16. **Citavi** exclusion reason: bibliographic management software.
17. **Comparative Toxicogenomics Database (CTD)** exclusion reason: it is a database, not a search strategy development tool.
18. **COVID-NMA** exclusion reason: it is a database, not a search strategy development tool.
19. **DisMaNET** exclusion reason: online version not available during review period. Download version last updated >2 years ago.
20. **Doctor Evidence** exclusion reason: search tool part of integrated review platform.
21. **Doctor Evidence DOC DATA** exclusion reason: search tool part of integrated review platform.
22. **Drug Herb Interaction** exclusion reason: it is a database, not a search strategy development tool.
23. **Drug-Gene Interaction** exclusion reason: it is a database, not a search strategy development tool.
24. **Epistemonikos** exclusion reason: it is a database, not a search strategy development tool.
25. **G-Bean** exclusion reason: last updated >2 years ago.
26. **GoPubMed** exclusion reason: last updated >2 years ago.
27. **GSscraper** exclusion reason: not a biomedical tool (search engine image scraper).
28. **Health Assessment Workspace Collaborative (HAWC)** exclusion reason: search tool part of integrated review platform.
29. **HelioBLAST** exclusion reason: it was not available during review period.
30. **Import.IO** exclusion reason: not a biomedical tool.
31. **JSTOR Text Analyzer** exclusion reason: it is a database, not a search strategy development tool.
32. **LaserAI** exclusion reason: search tool part of integrated review platform.
33. **Leaf** exclusion reason: it is a database, not a search strategy development tool.
34. **Lingo3G** exclusion reason: it is a fee-based tool.
35. **Litsearchr** exclusion reason: programming knowledge required to use it.
36. **L·OVE platform** exclusion reason: it is a database, not a search strategy development tool.
37. **Mapping MEDLINE** exclusion reason: last updated >2 years ago.
38. **MarkerDB** exclusion reason: it is a database, not a search strategy development tool.
39. **Medline (PubMed) Trend** exclusion reason: last updated >2 years ago.
40. **MedTerm Search Assistant** exclusion reason: last updated >2 years ago.
41. **MeSHSIM** exclusion reason: last updated >2 years ago.
42. **Metta** exclusion reason: last updated >2 years ago.
43. **NAILS (Network Analysis Interface for Literature Studies)** exclusion reason: last updated >2 years ago.
44. **Nested Knowledge** exclusion reason: search tool part of integrated review platform.
45. **NimbleMiner** exclusion reason: programming knowledge required to use it.
46. **NLM Medical Text Indexer (MTI)** exclusion reason: not a search tool.
47. **OmixLitMiner** exclusion reason: last updated >2 years ago.
48. **PaperBot** exclusion reason: last updated >2 years ago.
49. **Papers** exclusion reason: bibliographic management software.
50. **ParsCit** exclusion reason: last updated >2 years ago.
51. **Parsifal** exclusion reason: not a biomedical tool.
52. **PDQ-Evidence** exclusion reason: it is a database, not a search strategy development tool.
53. **PhenX Toolkit** exclusion reason: not a search strategy development tool.
54. **pitts.ai** exclusion reason: not a search strategy development tool.
55. **Publish or Perish** exclusion reason: not a search strategy development tool
56. **PubMed2XL** exclusion reason: bibliographic management software.
57. **PubVenn** exclusion reason: last updated >2 years ago.
58. **Qigga** exclusion reason: bibliographic management software.
59. **Qinsight** exclusion reason: no longer available.
60. **Quetzal** exclusion reason: no longer available.
61. **RAx** exclusion reason: search tool part of integrated review platform.
62. **RCT Tagger** exclusion reason: last updated >2 years ago.
63. **Researchr** exclusion reason: bibliographic management software.
64. **REviewER** exclusion reason: not a biomedical tool.
65. **RobotAnalyst** exclusion reason: not a search strategy development tool.
66. **Sample Size Search (SSS) Tool for PubMed** exclusion reason: no longer available.
67. **Science of Science (Sci2)** exclusion reason: last updated >2 years ago.
68. **Search Builder 1.0** exclusion reason: last updated >2 years ago.
69. **scite.ai** exclusion reason: not a search strategy development tool.
70. **Sebzer** exclusion reason: bibliographic management software.
71. **SensPrecOptimizer** exclusion reason: last updated >2 years ago.
72. **SESRA** exclusion reason: bibliographic management software.
73. **SLR.qub** exclusion reason: last updated >2 years ago.
74. **SRDB.PRO** exclusion reason: bibliographic management software.
75. **StArt Active Screener** exclusion reason: not a biomedical tool.
76. **Syras** exclusion reason: it is a fee-based tool.
77. **Sysrev** exclusion reason: it is a fee-based tool.
78. **Systematic Review Accelerator** exclusion reason: integrated tool (reviewed SRC search modules individually).
79. **Topictagger** exclusion reason: not a search tool.
80. **Trialstreamer** exclusion reason: it is a database, not a search strategy development tool.
81. **TRIP Rapid Reviews** exclusion reason: not a search tool.

